# Screening of Potential Biomarkers for Gastric Cancer with Diagnostic Value Using Label-free Global Proteome Analysis

**DOI:** 10.1101/2020.03.08.20031930

**Authors:** Yongxi Song, Jun Wang, Jingxu Sun, Xiaowan Chen, Jinxin Shi, Zhonghua Wu, Dehao Yu, Fei Zhang, Zhenning Wang

## Abstract

Gastric cancer (GC) is one of the most malignant tumors worldwide. Despite the recent decrease in mortality rates, the prognosis remains poor. Therefore, it is necessary to find novel biomarkers with early diagnostic value for GC. In this study, we present a large-scale proteomic analysis of 30 GC tissues and 30 matched healthy tissues using label-free global proteome profiling. Our results identified 537 differentially expressed proteins, including 280 upregulated and 257 downregulated proteins. The ingenuity pathway analysis (IPA) results indicated that the sirtuin signaling pathway was the most activated pathway in GC tissues whereas oxidative phosphorylation was the most inhibited. Moreover, the most activated molecular function was cellular movement, including tissue invasion by tumor cell lines. Based on IPA results, 15 hub proteins were screened. Using the receiver operating characteristic curve, most of hub proteins showed a high diagnostic power in distinguishing between tumors and healthy controls. A four-protein (ATP5B-ATP5O-NDUFB4-NDUFB8) diagnostic signature was built using a random forest model. The area under the curve (AUC) of this model was 0.996 and 0.886 for the training and testing set, respectively, suggesting that the four-protein signature has a high diagnostic power. This signature was further tested with independent datasets using plasma enzyme-linked immune sorbent assays, resulted in an AUC of 0.778 for distinguishing GC tissues from healthy controls, and using immunohistochemical tissue microarray analysis, resulting in an AUC of 0.805. In conclusion, this study identifies potential biomarkers and improves our understanding of the pathogenesis, providing novel therapeutic targets for GC.

## Introduction

Gastric cancer (GC) is one of the most common malignant, aggressive tumors, causing approximately 723,100 deaths worldwide in 2012 [1], particularly in East Asia [2]. It is a complex disease with histological and etiological heterogeneity [3]. Large genomic variations have been detected in GC patients [4]. Many patients are diagnosed with GC at a late stage due to the asymptomatic nature of the disease [5]. Despite a decrease in mortality rates in recent years, GC prognosis remains poor, with only 28.3% of new cases surviving for more than 5 years [6]. Our understanding of GC pathogenesis and molecular biology has improved, yet it is still necessary to identify novel biomarkers with early diagnostic value, to determine efficient diagnostic methods, and to discover new targets for treating GC.

Cancer development and progression requires molecular alterations at multiple levels including the genome, transcriptome, proteome, and metabolome [7]. In the past decade, numerous studies have examined molecular mechanisms of cancer using genomic and transcriptomic analyses. Protein dynamics are crucial for determining cancer phenotype, and the rapid development of quantitative proteomic approaches for studies on cancer proteomics stimulated investigations characterizing proteogenomic landscapes for many human cancers, including colorectal cancer [8], prostate cancer [9], breast cancer [10], lung adenocarcinoma [11], and ovarian cancer [12]. These efforts promoted the use of mass spectrometry (MS)-based proteogenomics for clinical use [13].

Several recent studies have investigated proteomic aspects of GC. Using the Isobaric Tags for Relative and Absolute Quantitation (iTRAQ) approach combined with high-resolution MS analysis, a previous study identified 3914 different proteins in six biopsies from different disease stages ranging from chronic gastritis and intestinal metaplasia to gastric adenocarcinoma [14]. Another study examined four GC tissues and four adjacent normal tissues and identified 431 differentially expressed protein (DEPs) using iTRAQ-based quantitative proteomic analysis [15]. This study showed correlations between MTA2 and HDAC1 expression levels and between lymph node metastasis and TNM staging for GC.

One major limitation of current GC proteomics studies is sample size. Small sample sizes introduce bias to findings and result in data inconsistencies. Also, due to individual heterogeneity, paired tumor and healthy control samples from the same patient should ideally be compared when searching for proteomic alterations [16]. Additionally, integrating proteomics and robust bioinformatics methods might help identify potential novel biomarkers with diagnostic power for GC.

In this study, we present a large-scale proteomic analysis of 30 GC tissues and 30 matched healthy tissues using label-free global proteome profiling. This proteomic analysis helped identify 537 DEPs, including 280 upregulated and 257 downregulated proteins. Results of the ingenuity pathway analysis (IPA) indicated that the sirtuin signaling pathway was most activated, whereas oxidative phosphorylation was the most inhibited pathway. Moreover, the most activated molecular function (MF) was cellular movement, including tissue invasion by tumor cell lines. Subsequently, 15 hub proteins were screened based on IPA enrichment results. Using the receiver operating characteristic (ROC) curve, most of these hub proteins had reliable diagnostic potential to distinguish between tumors and healthy controls. After that, a four-protein (ATP5B-ATP5O-NDUFB4-NDUFB8) diagnostic signature was built using a random forest model. The area under curve (AUC) of this model was 0.886 for the testing set, suggesting a high diagnostic potential. Additional independent datasets testing our four-protein signature using plasma enzyme-linked immune sorbent assays (ELISA) yielded an AUC of 0.778 and accuracy of 71.8% to distinguish GC from healthy controls, and visualizing protein expression with tissue microarray analysis yielded an AUC of 0.805. In conclusion, this study identified highly dysregulated proteins and potential biomarkers with potential use in detecting GC. These results further our understanding of GC pathogenesis and identify novel and specific therapeutic targets for this cancer.

## Results

### Global proteome profiling in the GC cohort

In this study, we conducted an integrated analysis of the global proteome profile of GC (**Figure 1**). Thirty primary tumor tissues and corresponding adjacent healthy tissues were obtained after surgical resection from a total of 30 GC patients at the First Hospital of China Medical University. The neoplastic purity analysis of all 60 samples is shown in Supplementary Figure 1A. High-resolution liquid chromatography-tandem MS (LC-MS/MS) was used to identify differences in proteomic profiles between the tumor and healthy samples. LC-MS/MS analyses were performed using MAXQUANT software (v1.4.1.2) [17]. The distribution of peptides, unique peptides, and identified proteins are shown in Supplementary Figure 1B. We used the label-free quantification (LFQ) algorithm embedded in MAXQUANT to quantify protein expression, and peptide-spectrum matching, false discovery rate (FDR), peptide FDR threshold, and protein FDR threshold were all set to 1%. A total of 10,615 proteins were identified in this study, with an average protein coverage rate of 28% (Supplementary Figure 2A and 2B). Of these, expression of 10,576 proteins was quantified, with expression of 2722 proteins quantified across all 30 tissue pairs (Supplementary Figure 2C).

**Figure 1.**
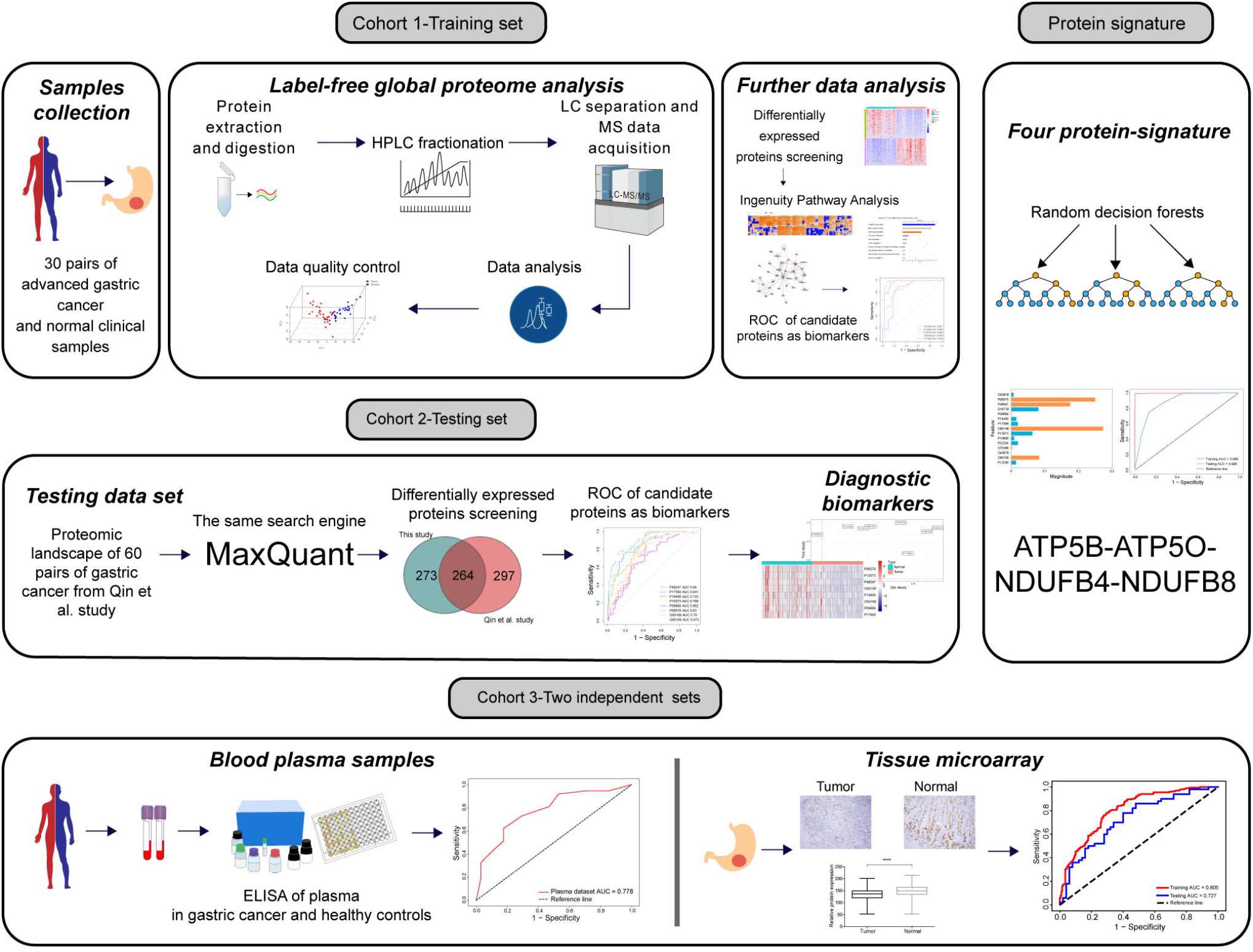
Workflow of the study. Screening of potential biomarkers for gastric cancer with diagnostic value using label-free global proteome analysis.

**Figure 2.**
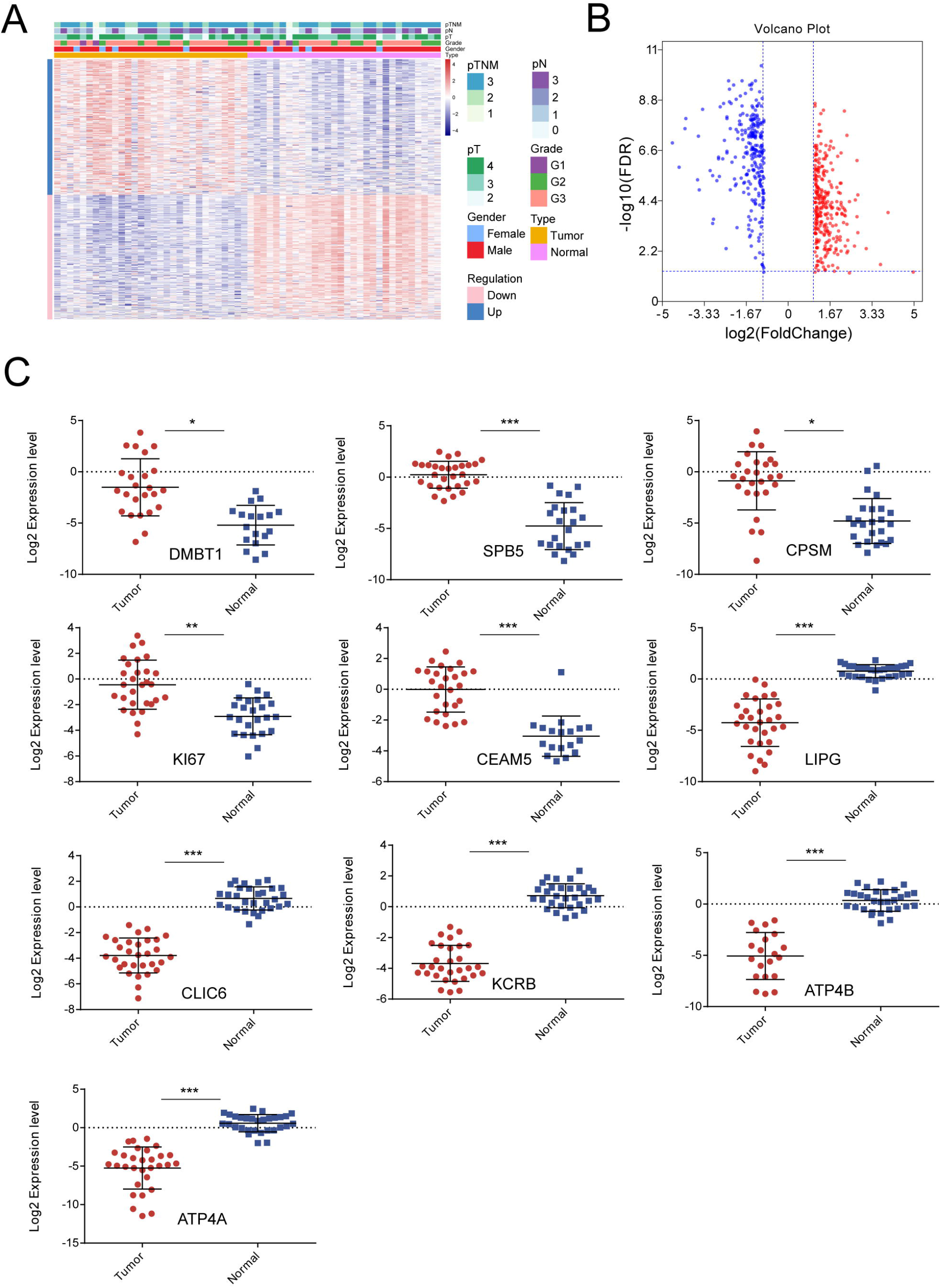
Proteomic features of differentially expressed proteins (DEPs) in gastric cancer (GC) **A**. Heatmap of DEPs in GC. pT: pathological Tumor stage, pN: pathological Lymph Node stage, pTNM: pathological TNM stage. **B**. A volcano plot for DEPs. The differential expression ratio of log2 (FC) (x axis) and the –log10 (FDR) value (y axis) were plotted for each identified protein. **C**. Expression profiles of the top ten significant DEPs in tumor and normal tissues. *P < 0.05, **P < 0.01, ***P < 0.001 (paired-samples *t*-test).

We began by calculating the ratio of tumor versus healthy tissues for one paired sample using LFQ values. After that, we generated Spearman’s correlation coefficient matrices for all 30 patients using protein ratios (Supplementary Figure 2D). The fraction of total (FOT) value was used to determine the distribution of protein expression profiles across all GC samples (Supplementary Figure 2E). Our results indicated consistent proteome identification and quantification throughout our study.

We next determined the coefficients of variation (CVs) and interquartile range (IQR) of the proteins. The overall CV decreased significantly when the FOT value was higher than 10^−5^ (Supplementary Figure 3A). Additionally, the increased performance of the IQR was discontinued when the FOT value was higher than 10^−5^ (Supplementary Figure 3B), suggesting that the most suitable value for accurate quantification was when the FOT value was 10^−5^. This result is consistent with the cut-off value from a previous GC proteomics study [16]. We also calculated the distribution of the quantitative samples and found a median CV of log10(FOT)+7. The median CV was significantly decreased when the number of quantitative samples was over 20 (Supplementary Figure 3C). Therefore, we filtered 3940 proteins using the above cut-off value and the number of quantitative samples (Supplementary Table 2). For these proteins, we used principal component analysis (PCA) to analyze expression in tumor tissues and corresponding matched healthy tissues (Supplementary Figure 3D). The proportion of variance of PC1, PC2, and PC3 was 19.06%, 6.91%, and 5.49%, respectively. These PCA results indicate a clear distinction between the proteomes of tumor and healthy tissues.

### Identification of DEPs in GC

We next assessed significant quantitative differences between tumor tissues and matched healthy tissues. DEPs were screened using a filter criterion of |log2-fold change (FC)|> 1 and FDR < 0.05. This identified 537 DEPs, including 280 upregulated and 257 downregulated proteins (**Figure 2A** and Supplementary Table 3). A volcano plot showing statistically significant DEPs between tumor and healthy tissues was constructed (**Figure 2B**). Expression levels of the top ten significant DEPs DMBT1, SPB5, CPSM, KI67, CEAM5, ATP4A, ATP4B, CLIC6, KCRB, and LIPG are shown in **Figure 2C** and **Table 1**. Subcellular localization analysis indicated that a large number of DEPs were annotated as mitochondrial, suggesting that the GC proteome is involved in tumor energy metabolism (**Figure 3A**). The gene ontology (GO) term enrichment analysis (Supplementary Table 4) revealed that upregulated DEPs in GC were significantly enriched in activities of the nucleolus, DNA, and RNA. Downregulated DEPs were mostly enriched in mitochondrial processes, cellular respiration, and oxidative phosphorylation (**Figure 3B**).

**Figure 3.**
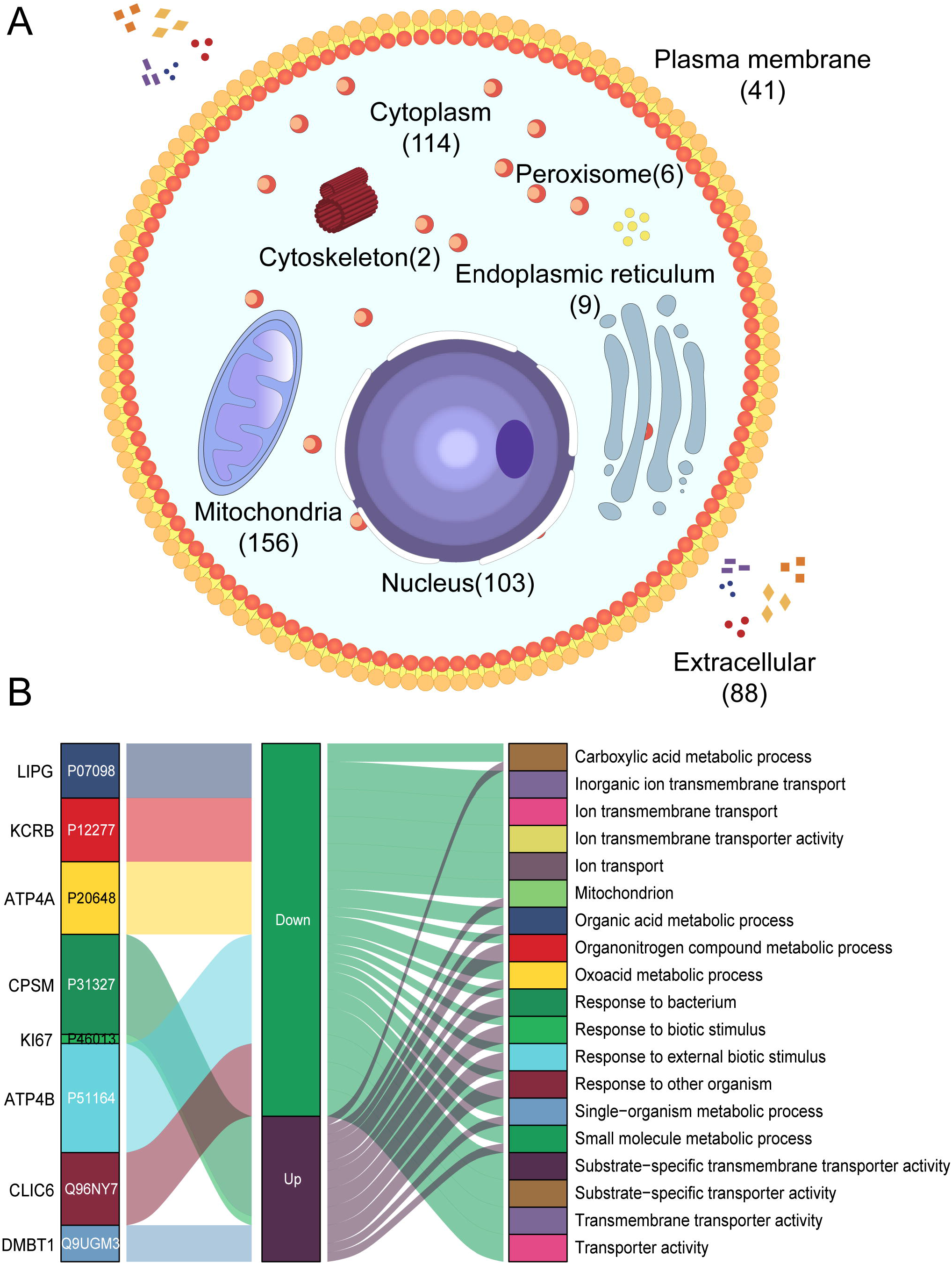
Subcellular distribution and GO term enrichment analysis of differentially expressed proteins (DEPs) in gastric cancer (GC) **A**. Subcellular distribution of DEPs. **B**. Graphical summary of DEPs and GO term enrichment analysis.

**Table 1.** The information of top 10 significant DEPs.

| UniProt ID | Gene Symbol | T-test p-value | FDR | Log2 T/N Ratio | Regulation |
| --- | --- | --- | --- | --- | --- |
| P20648 | ATP4A | 5.15E-09 | 1.02E-07 | -4.66 | down |
| P51164 | ATP4B | 1.53E-07 | 1.24E-06 | -4.37 | down |
| Q96NY7 | CLIC6 | 7.13E-10 | 2.55E-08 | -4.16 | down |
| P12277 | KCRB | 1.38E-10 | 8.61E-09 | -4.12 | down |
| P07098 | LIPG | 2.56E-15 | 1.01E-11 | -3.70 | down |
| Q9UGM3 | DMBT1 | 3.10E-02 | 4.48E-02 | 4.92 | up |
| P36952 | SPB5 | 3.78E-05 | 1.13E-04 | 3.92 | up |
| P31327 | CPSM | 1.33E-02 | 2.09E-02 | 3.62 | up |
| P46013 | KI67 | 2.39E-03 | 4.39E-03 | 3.05 | up |
| P06731 | CEAM5 | 3.76E-04 | 8.45E-04 | 2.71 | up |

### Proteomic pathways and potential hub proteins in GC

DEPs were further categorized using the IPA database to identify proteins with potential significant diagnostic values for GC. The pathway enrichment analysis indicated that DEPs were significantly enriched in oxidative phosphorylation, mitochondrial dysfunction, the sirtuin signaling pathway, and the tricarboxylic acid (TCA) cycle (Supplementary Table 5). Of the 25 significant signaling pathways, five pathways (sirtuin signaling, interferon signaling, inflammasome, IL-8 signaling, and neuroinflammation signaling) with z-scores > 0 were predicted to be activated in GC whereas 20 pathways (z-score < 0) were predicted to be inhibited (**Figure 4A**). The most inhibited pathways were oxidative phosphorylation and the TCA cycle. Of particular interest, oxidative phosphorylation is potentially inhibited and the sirtuin signaling pathway activated in GC (**Figure 4B**). All proteins identified to be involved in oxidative phosphorylation were downregulated.

**Figure 4.**
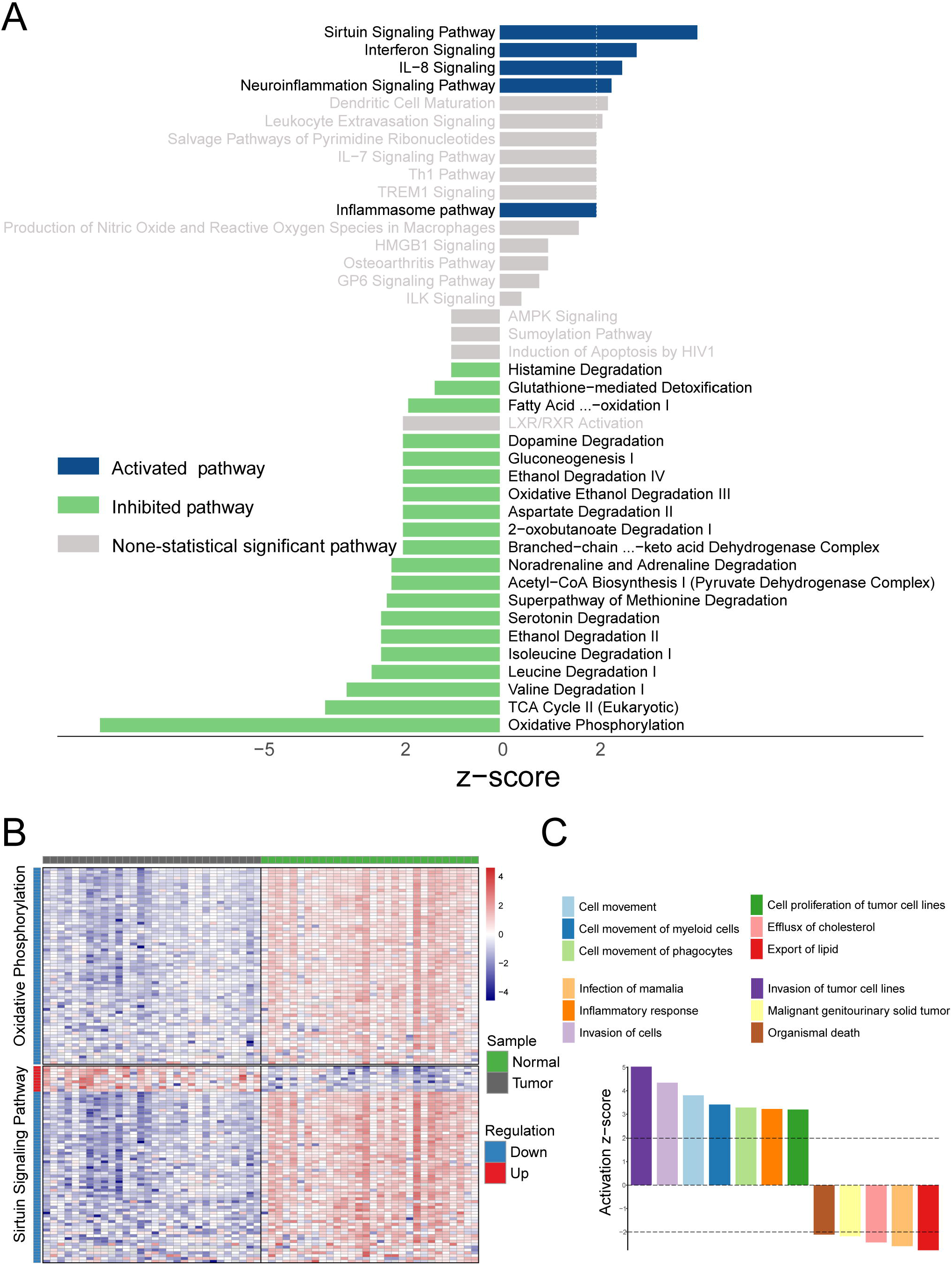
Activated or inhibited pathways and potential hub proteins in gastric cancer (GC) using ingenuity pathway analysis. **A**. Canonical pathway analysis of DEPs. **B**. Expression profiles of proteins in the oxidative phosphorylation pathway and the sirtuin signaling pathway. **C**. Disease and functional analysis for DEPs.

We performed disease and functional analyses for the above DEPs in IPA (**Figure 4C**), revealing that the most activated function was cellular movement, including tissue invasion by various tumor cell lines. Tumor cell line proliferation, adhesion, and inflammatory responses were also found to be activated in tumors. The most inhibited function was lipid export. The network associated with regulation of tissue invasion by tumor cell lines and lipid export was connected by five proteins: ACAT1, CAV1, CTSS, S100A12, and S100A9 (**Figure 5A**).

**Figure 5.**
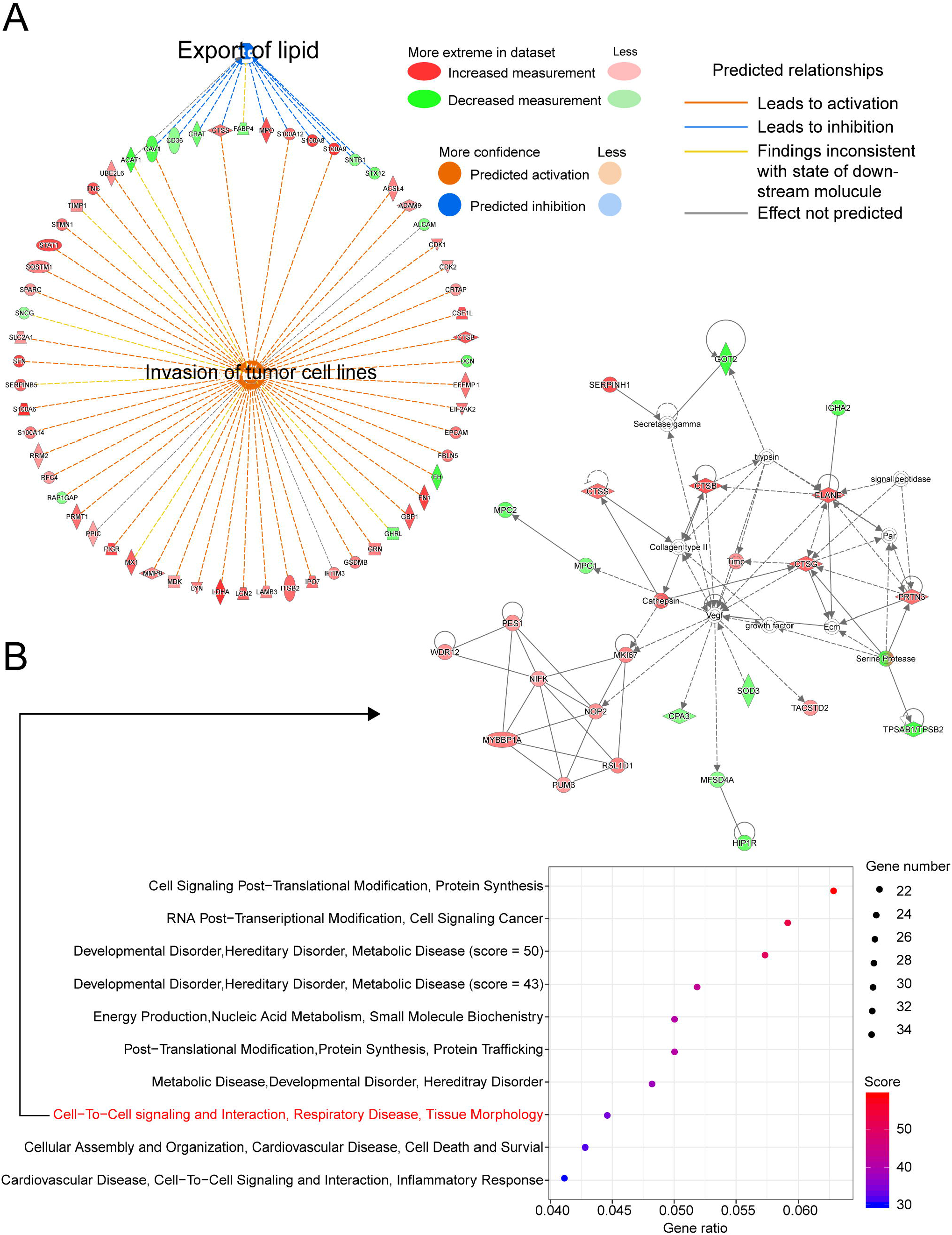
Interaction network and biomarker analysis for DEPs. **A.** Interaction network analysis of tissue invasion by tumor cell lines and lipid export. **B.** Biomarker analysis for DEPs.

Our biomarker analysis identified 72 potential biomarkers associated with cancer and gastrointestinal diseases, and the network analysis implicated 25 networks. Significant regulatory networks with scores > 30 were found associated with cell signaling, post-translational modification, protein synthesis, protein trafficking, energy production, nucleic acid metabolism, small molecule biochemistry, and cell-to-cell signaling and interaction (**Figure 5B**).

### Screening potential diagnostic markers in GC

We next screened fifteen hub proteins from the enriched canonical pathways, biomarker analyses, and the top ten protein-protein interaction networks (Supplementary Table 6). The diagnostic performance of each protein was assessed using the ROC curve. Most proteins (13 out of 15) showed a high AUC (> 0.80) between GC and healthy tissues, suggesting that these hub proteins might have discriminating potential as GC diagnostic markers (**Figure 6A**). Among these, NDUB8 (O95169) and CX7A2 (P14406) had AUC of 0.98 and 0.978, respectively. The 95 % confidence interval (CI) of the AUC and p-values compared with the reference line are shown in Supplementary Table 7.

**Figure 6.**
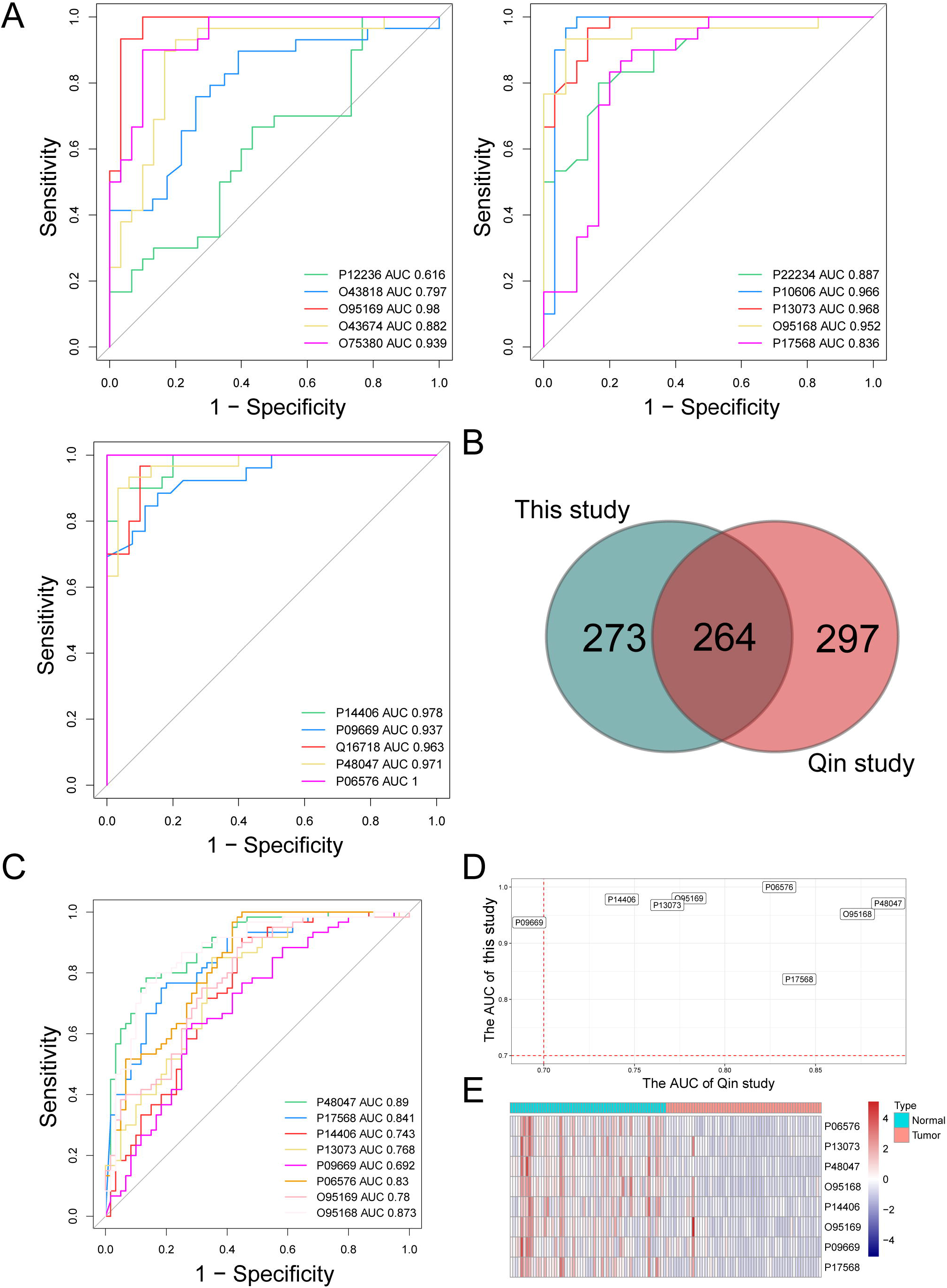
Screening and validation of potential proteins as diagnostic markers for gastric cancer in our and testing datasets. **A**. ROC curve for each hub protein in the training phase. **B**. Venn diagram summarizing the number of DEPs between our study and the testing phase. **C**. ROC curve for each shared hub protein in the testing phase. **D**. Identification of potential and independent diagnostic proteins as biomarkers. **E**. Expression profile of significant shared hub proteins in the testing phase.

To test our newly identified proteins, we used an independently published cohort of 60 GC samples and matched healthy samples [16]. A total of 561 DEPs including 345 upregulated and 216 downregulated proteins were selected using the same filter criteria (Supplementary Table 8). When compared with the results of our initial cohort (**Figure 6B**), 264 DEPs were shared between the two datasets, and 8 of the 15 hub proteins discussed above were differentially expressed in the testing phase. As shown in **Figure 6C**, ATP5O (P48047, AUC = 0.89) and NDUB4 (O95168, AUC = 0.873) were the two proteins with the highest predictive power. Proteins with AUC > 0.70 were considered potential independent diagnostic biomarkers (**Figure 6D**). Only one protein (P09669) was excluded. Expression profiles of significant hub proteins found in the testing phase are shown in **Figure 6E**.

### Establishment and validation of a four-protein signature

Although the above results indicate that single proteins may hold significant discriminating potential, we investigated the possibility of building a multi-protein signature with increased diagnostic potential, sensitivity, and specificity. We used a random forest model (**Figure 7A**) including the performance of each protein. The best-performing proteins were found to be NDUB4 (O95168), ATPB (P06576), ATPO (P48047), and NDUB8 (O95169) (**Figure 7B**). After increment feature selection, AUC, sensitivity, specificity, and accuracy were found to be constant and suitable when the four-protein signature was built (**Figure 7C**). The AUC of this protein signature in the training set was 0.996 (**Figure 7D**, red line) and the accuracy was 98.3 %. Thus, we built a four-protein signature (ATP5B-ATP5O-NDUFB4-NDUFB8) with high diagnostic potential for GC.

**Figure 7.**
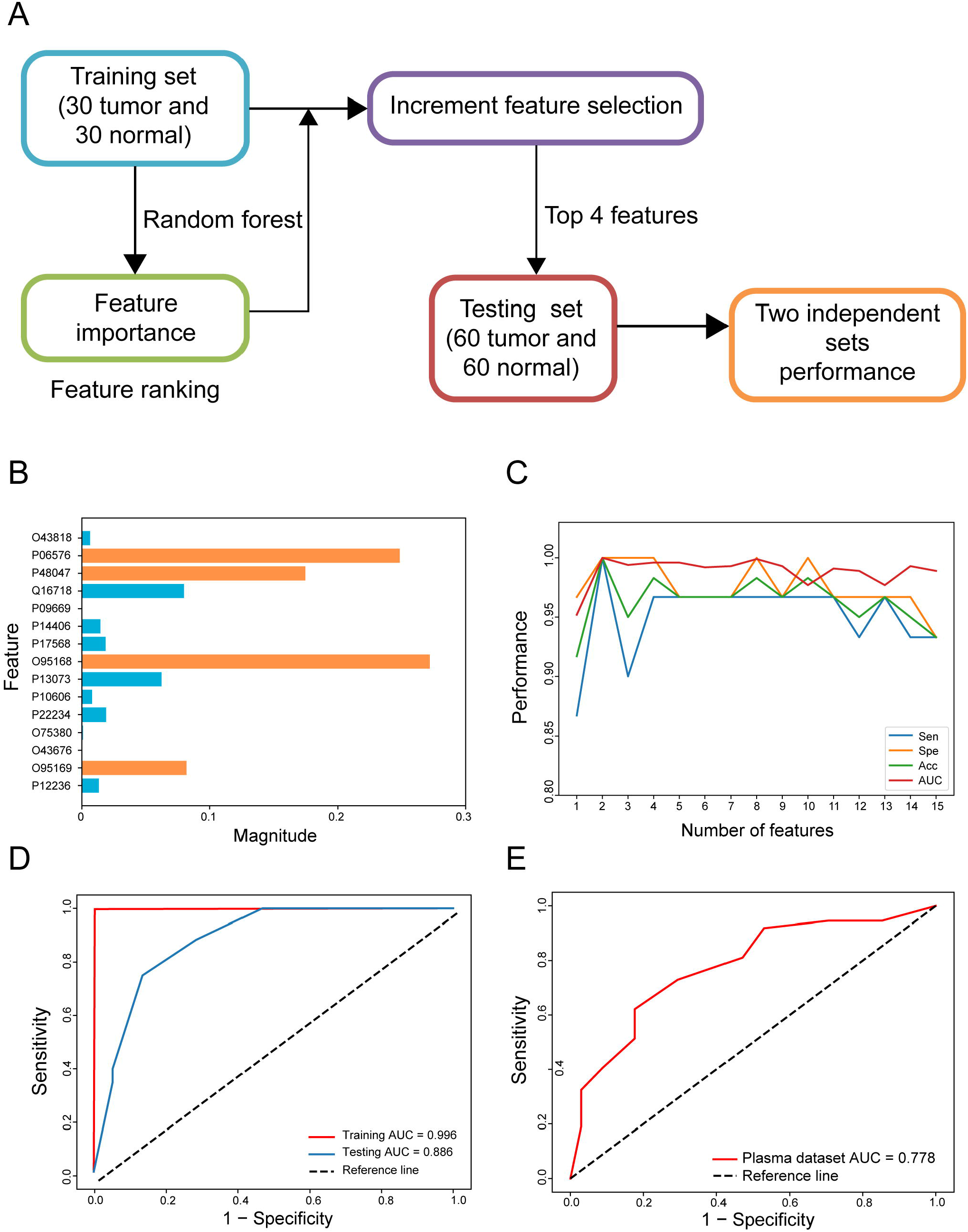
Establishment and validation of a four-protein signature for the diagnosis of gastric cancer. **A**. Design of the random forest model. **B**. Feature performance of each protein in the random forest model. **C**. Increment feature selection. AUC, sensitivity, specificity, and accuracy became constant and suitable when the four-protein signature was built. Sen: sensitivity, Spe: specificity, Acc: accuracy. **D**. ROC curve for the four-protein diagnostic signature for the training set (red line) and testing set (blue line). **E**. ROC curve for the four-protein diagnostic signature for the plasma samples.

To test the stability and diagnostic power of the four-protein signature, we analyzed GC and healthy tissues using LC-MS/MS, a testing cohort consisting of proteomic dataset from 60 GC patients [16] using label-free analysis. The AUC of the four-protein signature was 0.886 and its accuracy was 80.0 %, suggesting that this protein panel had a high diagnostic value for GC (**Figure 7D**, blue line).

The above results were obtained from tumor and healthy tissues collected from patients and analyzed using LC-MS/MS. To further extend our study for clinical use and noninvasive detection, we used blood plasma samples from GC patients and healthy individuals to evaluate the potential diagnostic value of the protein signature for detecting GC using blood. We performed ELISA on plasma samples from 37 GC patients and 34 healthy controls for the validation phase. Our results showed that the protein signature had an AUC of 0.778 and accuracy of 71.8 % to distinguish GC tissues from healthy controls (**Figure 7E**).

Finally, we tested 251 pairs of GC tissues using tissue microarray. As shown in **Figure 8A-D**, we first determined differential expression patterns for the four proteins by immunohistochemical staining on tumor and normal tissues (p-values < 0.05). As this dataset included an adequate number of samples, we could divide the samples into internal training (80% samples) and testing (20% samples) sets. The AUC for the internal training set was 0.805, with an AUC of 0.727 for the internal testing set (**Figure 8E)**. The p-values compared with reference line for all datasets are shown in Supplementary Table 9.

**Figure 8.**
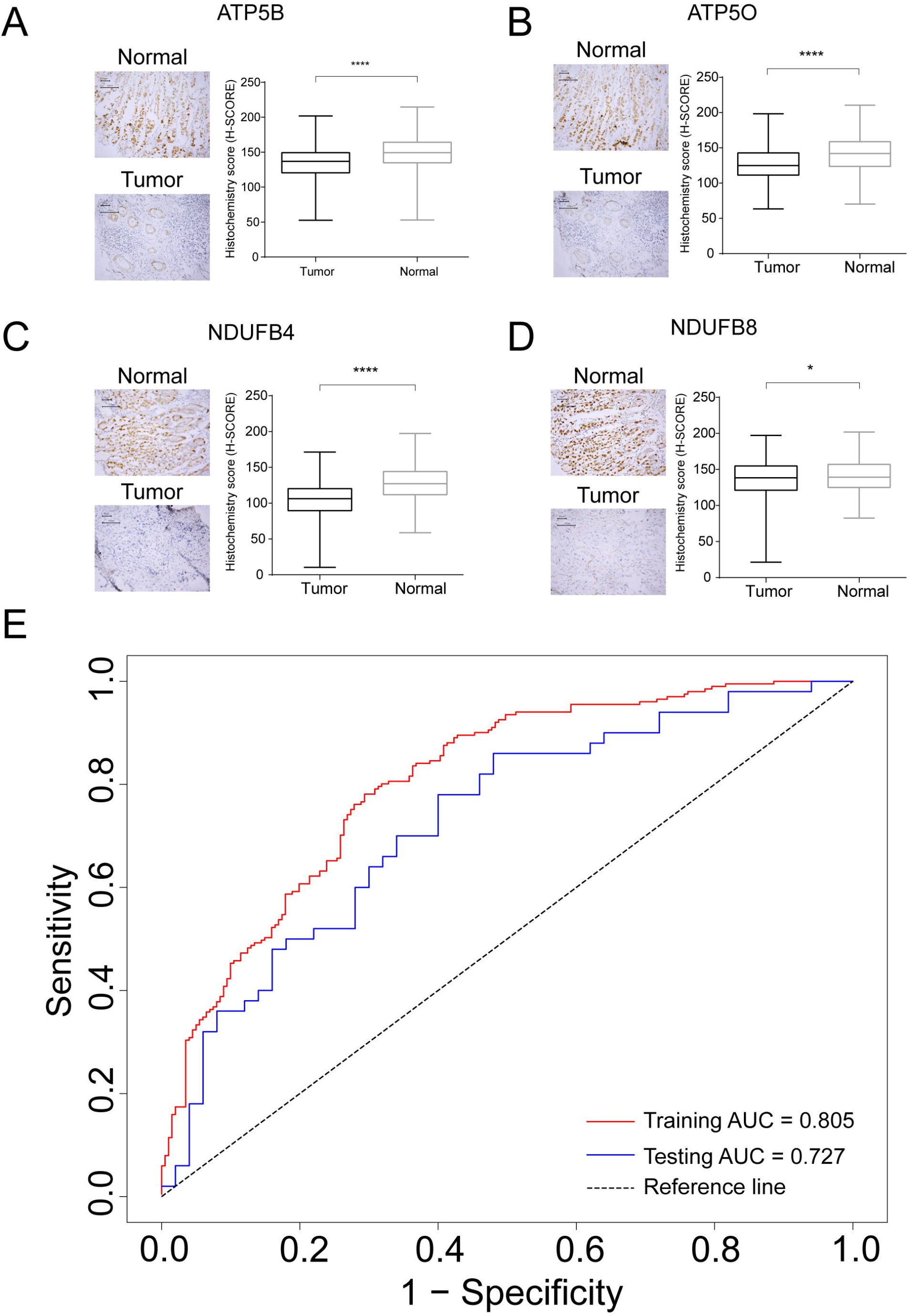
Validation of a four-protein signature for the diagnosis of gastric cancer in tissue microarray. The immunohistochemical staining of **A**. ATP5B, **B**. ATP5O, **C**. NDUFB4, and **D**. NDUFB8 in tumor and normal GC tissues. Scale bars represent 50 and 100 μm. Each case was at ×200 magnification. *P < 0.05, **P < 0.01, ***P < 0.001, ****P < 0.0001 (paired-samples *t*-test). **E**. ROC curve for the four-protein diagnostic signature for the tissue microarray.

At the same time, we tested the AUC values of each individual protein from the four-protein combination in above datasets. We found very small AUC values (Supplementary Figure 4). In sum, our results suggest that this novel four-protein signature (ATP5B-ATP5O-NDUFB4-NDUFB8) has a high diagnostic power for GC.

## Discussion

In this study, we present a large-scale proteomic analysis of GC using label-free global proteome profiling based on 30 GC tissues paired with 30 healthy tissues. A four-protein (ATP5B-ATP5O-NDUFB4-NDUFB8) diagnostic signature was built using a random forest model. Our findings might help to understand the pathogenesis of GC and provide novel and specific therapeutic targets for this disease.

### Biomarkers and proteomics in cancer

Biomarkers may play important roles in the diagnosis and treatment of cancers by enabling early detection and diagnosis [18]. Furthermore, the rapid development of quantitative proteomic approaches has allowed researchers to analyze biomarkers for human tumors. For instance, proteomics were used for biomarker discovery for colorectal cancer, including the role of protein phosphorylation and cancer stem cells [19]. Clinical proteomics has also proven to be a promising tool for improving personalized medicine for colorectal cancer using blood, stool, and biopsy samples [20]. Moreover, MS-based proteomics have been used for drug discovery and development [21].

Other groups have also performed proteomic studies for GC. Huang et al. performed a quantitative proteomic study using ten GC serum samples and healthy controls using tandem mass tags [22], and identified 594 serum DEPs with a cut-off value of 1.2-FC. The DEPs 1C12, PIGR, S10A8, AOC3, FHL1, GGCT, NCAM1, and SYNEM were also identified in our study. Liu et al. performed label-free LC-MS/MS using GC tissues and healthy tissues from six patients [23] and found 87 DEPs. Of these 87 proteins, ATPB, ATP4B, NDUB9, and NDUAD were also identified in our study. However, we used a more stringent screening criteria with a cut-off value of 2-fold change and an FDR < 0.05. The analysis of our testing set (60 pairs) revealed that 264 DEPs were shared between the two datasets. The DEPs identified by us are different from those of previous studies, likely due to the different platforms, quantitative methods, and screening criteria used. Therefore, more accurate analytical methods and a larger number of samples are necessary to confirm our findings.

Diagnostic test performance is often assessed by measuring ROC, AUC, sensitivity, and specificity. For an example, a label-free quantitative proteomic study was performed to diagnose periodontal diseases using saliva [24], and the authors found that 12 proteins presented the highest AUC (AUC 0.83-0.91) between healthy and diseased tissues. Yang et al. used targeted proteomics coupled with immunoaffinity enrichment to investigate epithelial ovarian cancer samples and found that the combined AUC for serum carbohydrate antigen 125 (CA125) and heat shock protein 27 was 0.88, which was significantly higher than that of CA125 alone [25]. Jiang et al. performed iTRAQ labeling and LC-ESI-MS/MS to evaluate a discovery group of four GC samples and four adjacent healthy tissues [26], and found an AUC of 0.734 for GLS1 and GGCT co-expression, suggesting that the level of co-expression had high clinical value as a diagnostic biomarker for early GC.

### IPA pathway analysis

Our results highlighted five pathways (sirtuin signaling, interferon signaling, inflammasome, IL −8 signaling, and neuroinflammation signaling) with z-scores > 0 predicted to be activated in GC. Among these, the sirtuin signaling pathway was the most activated. Sirtuins are members of the class III histone deacetylase family of enzymes [27], and mammalian sirtuins are classified into seven groups (SIRT1-7) [28]. Sirtuin signaling pathways regulate stem cell functions crucial for normal embryonic development and adult tissue homeostasis [29]. In cancer, sirtuins are implicated in producing cancer cells capable of self-renewal and differentiation, resulting in tumor growth. Several studies also confirmed the role of sirtuins in stimulating epithelial-mesenchymal transition [30, 31]. The interferon signaling pathway and other innate immune signaling pathways in tumor cells were shown to be determinants of treatment response and resistance [32]. These pathways may serve as an alternative immunotherapeutic strategy in GC [33, 34], and our results suggest that these activated pathways may play important roles in the pathogenesis of GC.

### TCA cycle and oxidative phosphorylation

The most inhibited pathways in GC were oxidative phosphorylation and the TCA cycle. This is consistent with another report on GC [14]. Recent studies have shown that cancer cells exhibit significant metabolic changes in mitochondrial dynamics and function [35]. Tumor growth is regulated by the TCA cycle and oxidative phosphorylation in mitochondria. According to the Warburg Effect [36], cancer cells are likely to increase energy metabolism via aerobic glycolysis rather than oxidative phosphorylation. Therefore, the regulation of energy metabolism via oxidative phosphorylation and the TCA cycle tends to be inhibited in GC, with aerobic glycolysis stimulated.

### The four proteins of the GC signature

Our study identifies a four-protein signature with diagnostic value for GC (ATP5B-ATP5O-NDUFB4-NDUFB8). Of these, ATP5B has not been reported upon in previous studies on GC, however, it was found to be downregulated in clear cell renal cell carcinoma [37]. In glioblastoma, ATP5B mRNA levels were significantly higher in tumor cells compared with healthy brain blood vessels, and microvascular proliferation was significantly higher [38]. ATP5O was reported as one member of 8 mitochondrial proteins (NDUFS5, VDAC3, ATP5O, IMMT, MRPL28, COX5B, MRPL52, PRKDC) which generated a compact gastric mitochondrial gene signature for predicting tumor progression and overall survival of GC patients [39]. ATP5O Gene expression was also downregulated in clear cell renal cell carcinoma [37]. NDUFB8 was found to be hypermethylated in glioblastoma [40]. However, NDUFB4 has been rarely reported upon in human cancers.

ATP5B and ATP5O are ATP synthases, and NDUFB4 and NDUFB8 are two members of the NADH dehydrogenase (ubiquinone) 1 beta sub-complex. Mitochondrial dysfunction is common in cancer, and mitochondrial electron transport chains are often affected in carcinogenesis [41, 42]. Mitochondrial dysfunction is involved in cancer cell metabolism, apoptosis and autophagy. Using antibiotics as anticancer drugs has been considered as potential anticancer therapy [43]. Therefore, targeting mitochondrial alterations might be a promising strategy for the development of tools for GC diagnosis, prognosis and treatment.

### The performance of protein signature in plasma

The performance of our protein signature was found to be better when measured in tissues (AUC = 0.996 and 0.886) than in plasma (AUC = 0.778). A major reason for this might be due to the stability of different experimental tests including LC/MS-MS and ELISA. A second reason might result from the complexity of the multi-step process for analyzing blood plasma samples from tumor and healthy controls. However, the blood plasma test is still of value. Moreover, as described above, our protein signature closely associated with energy metabolism. Therefore, this diagnostic protein signature may be applicable to other cancers in addition to GC.

### The differences between our and Qin’s study

Recently, Qin’s group presented a dataset providing information on proteomics of gene products and mutations in cancer driver genes from 84 diffuse-type GC patients [16]. They divided the patients into three molecular subtypes that provided a wealth of information on diffuse GC signaling pathways and demonstrated the benefits of proteomic analysis in cancer molecular subtypes. In contrast to with Qin’s study, our project focuses on mass spectrometry-based proteomics and bioinformatic algorithms to screen biomarkers and models with diagnostic value in GC. In our study, we identified hub proteins with high diagnostic power in distinguishing tumors and normal controls. We further built a four-protein (ATP5B-ATP5O-NDUFB4-NDUFB8) diagnostic signature using a random forest model then verified it GC tissues as well as with two independent datasets of plasma and tissue microarrays. Our study identified potential biomarkers and may help increase our understanding of GC pathogenesis and provide novel and specific diagnostic targets for this cancer.

As shown in our multiple rounds of testing, our narrowed-down four-protein signature had high diagnostic power between tumor tissues and healthy controls, suggesting its potential use as a novel clinical biomarker for GC. However, further large-scale validation studies are necessary to confirm this finding.

### Conclusion

In summary, this study increased our understanding of GC pathogenesis and identified potential biomarkers to provide novel and specific therapeutic targets for this cancer.

## Materials and Methods

### Selection of specimens and clinical information

Thirty tumor specimens and 30 matched healthy tissues were obtained from 30 GC patients following surgical resection at the First Hospital of China Medical University (Shenyang, China). Tissue sections were stained with hematoxylin and eosin to evaluate tumor purity by a certified pathologist in the hospital. Tumor samples consisting of at least 60% cancer cells were retained for further analysis (Supplementary Figure 1A and Supplementary Table 1). Each sample was collected within 30 min after surgical resection, cleaned, transferred to sterile freezing tubes, and cryopreserved in liquid nitrogen until further use. All samples were staged according to the seventh edition of The American Joint Committee on Cancer staging system.

In the testing phase, the diagnostic value of candidate proteins was estimated using the plasma of 71 individuals (37 GC patients and 34 healthy controls). In brief, 2 mL of whole peripheral venous blood were collected from each sample and transferred to a purple-top EDTA tube. Plasma samples were separated following a two-step centrifugation protocol (3000 rpm for 5 min at 4 °C, and 12,000 rpm for 15 min at 4 °C) within 4 h after collection, then samples were transferred to RNase/DNase-free tubes and stored at –80 °C until further use.

This study was conducted according to the principles established by the Declaration of Helsinki and was approved by the Human Research Ethics Committee of China Medical University (Shenyang, China). A written informed consent was obtained from all patients.

### Protein extraction and tryptic digestion

GC tissues were ground into powder in liquid nitrogen and suspended in an ice-cold lysis buffer [8 M urea (V900119-500G, VETEC, China), 5 mM dithiothreitol (DTT, D9163-5G, SIGMA, Canada), 1 % (v/v) protease inhibitor cocktail (539134, Merck, Darmstadt, Germany), 3 μM trichostatin A (TSA, V900931-5MG, VETEC, China), 50 mM nicotinamide (NAM, N0636-500G, SIGMA, USA)] with occasional sonication. Cell lysates were centrifuged at 12,000 ×g at 4 °C for 10 min, and the resulting supernatants were collected. Total protein concentration was determined from lysates using the 2-D Quant kit (80-6483-56, GE Healthcare, USA) according to the manufacturer’s instructions. Proteins were then precipitated with 15 % trichloroacetic acid (TCA, T4885-1KG, SIGMA, China) for 4 h at 4 °C, and the resulting precipitate was washed three times with cold (–20 °C) acetone. Dried protein pellets were resuspended in 100 mM tetraethylammonium bromide (TEAB, T7408-500ML, SIGMA, Switzerland) and digested with trypsin (V5111, Promega, USA) at an enzyme-to-substrate ratio of 1:50 for 12 h at 37 °C. Peptides were reduced with DTT and alkylated with iodoacetamide (V900335-5G, VETEC, China) in the dark. Complete digestion was ensured by performing a second digestion with trypsin at an enzyme-to-substrate ratio of 1:100 for 4 h at 37 °C.

### Peptide fractionation using high performance liquid chromatography (HPLC)

Samples were fractionated using high pH reverse-phase HPLC using an Agilent 300Extend C18 column (particle size, 5 μm; ID, 4.6 mm; length, 250 mm. Agilent Technologies, USA). Briefly, peptides were first separated into 80 fractions using a gradient of 2 % to 60 % acetonitrile in 10 mM ammonium bicarbonate pH 10 for 80 min, then they were pooled into multiple fractions (ten for label-free proteome) and dried using vacuum centrifugation.

### Liquid chromatography-tandem MS analysis

For label-free proteomic analysis, peptides were dissolved in 0.1% formic acid (FA, 94318-50ML-F, Fluka, USA) and loaded onto a reversed-phase pre-columns (Acclaim PepMap 100, Thermo Scientific). Peptides were separated using a reversed-phase analytical column (Acclaim PepMap RSLC, Thermo Scientific, USA). The elution gradient used was 5 % to 10 % of solvent B (0.1 % FA in 98 % ACN) for 4 min; 10 % to 23 % of solvent B for 36 min; 23 % to 35 % of solvent B for 12 min; 35 % to 80 % of solvent B for 4 min; and 80 % of solvent B for 4 min. Elution was performed at a constant flow rate of 300 nl/min using an EASY-nLC 1000 ultra-performance liquid chromatography (UPLC) system. Peptides subjected to post-translational modifications were separated in a similar way and analyzed using a Q Exactive TM Plus hybrid quadrupole-Orbitrap mass spectrometer (ThermoFisher Scientific, USA).

Peptides were subjected to a nanoelectrospray ionization source followed by tandem mass spectrometry (MS/MS) using a Q Exactive TM Plus (Thermo) mass spectrometer coupled online to a UPLC system. Intact peptides were detected using Orbitrap at a resolution of 70,000. Peptides were selected for MS/MS using a normalized collision energy of 30; ion fragments were detected using Orbitrap at a resolution of 17,500. A data-dependent procedure that alternated between one MS scan followed by 20 MS/MS scans was used for the top 20 precursor ions above a threshold ion count in the MS survey scan with different dynamic exclusion. For proteome detection, the ion count threshold and dynamic exclusion used were 1E4 and 30.0 s, respectively.

### Analysis of global proteomics data

The resulting MS/MS data were processed using MaxQuant with an integrated Andromeda search engine (version 1.4.1.2). Tandem mass spectra were searched against a SwissProt human database (downloaded on August 2015) concatenated with a reverse decoy database. Trypsin/P was defined as the cleavage enzyme allowing up to two missing cleavages. For proteomic analysis, the first search range was set to 5 ppm for precursor ions, and the main search range was set to 5 ppm and 0.02 Da for fragment ions. The carbamidomethylation of cysteines was defined as the fixed modification, and the oxidation on methionine was defined as the variable modification. The quantification method used was LFQ, the FDR was adjusted to < 1 %, and the minimum score for modified peptides was > 40.

### Identification of DEPs

To identify the proteins differentially expressed between tumor and healthy tissues, DEPs were defined as meeting the following criteria: |log2-Fold change (FC)|> 1 and FDR < 0.05. The heatmap of expression profiles was drawn using the *pheatmap* package in R language. GO enrichment analysis was performed to assess the functional biological role of DEPs. For GO term enrichment, including MF, biological process, and cellular component, a two-tailed Fisher’s exact test was used to determine the enrichment of DEPs against all proteins identified in-house using a Perl script. Categories with an FDR < 0.05 were considered significant. The graphical summary of GO results was drawn using the *ggalluvial* R package.

### IPA

The molecular and biological functions of DEPs were analyzed using IPA (Qiagen, https://www.qiagenbioinformatics.com/products/ingenuity-pathway-analysis/) [44]. This analytical tool includes canonical pathway analysis, interaction network analysis, disease and functional analysis, and a biomarker filter. The two statistical indicators of IPA used were the p-value and z-score. P-values < 0.05 were considered statistically significant, and the z-score was calculated using an internal algorithm and IPA standard. The molecular interaction was activated when the z-score was > 0 and inhibited when the z-score was < 0.

### Screening and validation of diagnostic markers using ROC analysis

The ROC curve was used to further select potential biomarkers with diagnostic power by determining the specificity and sensitivity of each protein, and the AUC was used to estimate the diagnostic value. The ROC curve was drawn using the *pROC* R package.

For the testing cohort, a proteomic dataset from 60 GC patients obtained using label-free analysis was used [16]. Raw data of all samples were downloaded, and the same search engine MaxQuant as well as the same filter criteria were used to identify DEPs. Based on these results, ROC analysis was conducted to test the diagnostic power of potential hub proteins in the testing cohort.

### Analysis using a random forest model

To further build a multi-protein signature with diagnostic power, the significant hub DEPs were used as attributes, and an analysis using a random forest model was performed in-house using a python script. In brief, this method was used together with 10-fold cross-validation for the training set to build a four-protein signature. Moreover, the diagnostic value of this model was verified using ROC analysis. Sensitivity, specificity, accuracy, and AUC were used to determine predictive values.

In the testing phase, based on the expression of these four proteins, an analysis using a random forest model was performed to prove the accuracy of the diagnostic value of the protein signature.

### Validation of the four-protein signature using ELISA

The diagnostic power of the four-protein signature was validated using 71 plasma samples (37 from GC patients and 34 from healthy controls). In brief, human plasma ATP synthase, H+ transporting, mitochondrial F1 complex, beta polypeptide (ATP5B), ATP synthase, H+ transporting, mitochondrial F1 complex, O subunit (ATP5O), NADH: ubiquinone oxidoreductase subunit B4 (NDUFB4), and NADH: ubiquinone oxidoreductase subunit B8 (NDUFB8) levels were measured using commercial ELISA kits (Catalog No. JL46945 for ATP5O, JL46944 for ATP5B, JL47136 for NDUFB4, JL47139 for NDUFB8, Shanghai Jianglai Biotech, Shanghai, China) according to the manufacturer’s protocols.

### Validation of the four-protein signature using tissue microarray

Immunohistochemistry was performed using 5 µm thick TMA sections as previously described [45]. After dewaxing in xylene and rehydration through graded alcohol, sections were placed in 3 % hydrogen peroxide to block endogenous peroxidase. For antigen retrieval, sections were boiled in citrate/EDTA buffer (citrate buffer for ATP5O, ATP5B and NDUFB4, Catalog No. ZLI-9065, ZSGB-BIO, Beijing, China; EDTA buffer for NDUFB8, Catalog No. R20904, Shanghai Yuanye Bio-Technology, Shanghai,China). After blocking with goat serum (Catalog No. SP-9001 for ATP5B and NDUFB8, ZSGB-BIO, Beijing, China; Catalog No. SP-9002 for ATP5O and NDUFB4, ZSGB-BIO, Beijing, China), sections were incubated overnight with anti-ATP5O/anti-ATP5B/anti-NDUFB4/anti-NDUFB8 antibodies (anti-ATP5O: Catalog No. ab110276, diluted 1:5000, Abcam, Cambridge, UK; anti-ATP5B: Catalog No.33031, diluted 1:5000, SAB, College Park, Maryland, USA; anti-NDUFB4: Catalog No. ab110243, diluted 1:1000, Abcam, Cambridge, UK; anti-NDUFB8: Catalog No. ab192878, diluted 1:8000, Abcam, Cambridge, UK) at 4 °C. Sections were then further incubated with reagents from the SP IHC Kit (Catalog No. SP-9001 for ATP5B and NDUFB8, ZSGB-BIO, Beijing, China; Catalog No. SP-9002 for ATP5O and NDUFB4, ZSGB-BIO, Beijing, China), 15 min for each reagent. Sections were visualized using DAB (Catalog No. ZLI-9010, ZSGB-BIO, Beijing, China) and Hematoxylin.

Slides were analysed with Panoramic MIDI digital scanner (3DHISTECH, Budapest, Hungary). Quantitative image analysis was performed by the Quant center software using histochemistry score (H-SCORE).

### Statistical analysis

Statistical analyses were performed using the SPSS software version 19.0 (SPSS, Inc., Chicago, IL, USA). The student’s *t*-test was used to evaluate differences between DEP expression in GC tissues and in matched healthy tissues. ROC and AUC were used to assess the diagnostic value of each candidate protein and protein signature. Two-tailed p-values < 0.05 were considered statistically significant.

## Data Availability

The MS proteomics data were deposited in the iProX database at https://www.iprox.org/ with the dataset identifier IPX0001590000. The raw dataset is free for all users.

## Authors’ contributions

YXS and JW designed experiments. JXS (Jingxu Sun), XWC, DHY and FZ contributed to the literature review. JXS (Jinxin Shi) and ZHW collected the clinical gastric cancer samples. YXS and JW helped to perform experiments. JW wrote the initial draft of the manuscript. ZNW designed and supervised the study, developed the concept and edited the paper. All authors have approved the final version of the manuscript.

## Acknowledgements

We thank the department of Surgical Oncology of The First Hospital of China Medical University for providing human gastric tissue samples and plasma samples. We thank Jingjie PTM BioLab (Hangzhou, China) for technical assistance. We also thank Elsevier (https://webshop.elsevier.com/language-editing-services/) for language editing services.

## Supplementary Material

### Supplementary Tables Legends

**Supplementary Table 1** The clinical information across all AGC samples in this study.

**Supplementary Table 2** Filtered proteins used in further analysis.

**Supplementary Table 3** Identification of differentially expressed proteins in AGC by proteomic analysis.

**Supplementary Table 4** GO term enrichments analysis of DEPs.

**Supplementary Table 5** IPA canonical pathway analysis of DEPs

**Supplementary Table 6** 15 potential hub proteins by ingenuity pathway analysis.

**Supplementary Table 7** The 95% confidence interval (CI) of AUC and p-value compared with reference line in this study and Qin’s study.

**Supplementary Table 8** Identification of differentially expressed proteins in testing phase (Qin’s study).

**Supplementary Table 9** The p-value compared with reference line in all datasets.

### Supplementary Figure Legends

**Supplementary Figure 1 Quality control of clinical samples and global proteome profiling**

**A**. The neoplastic purity analysis of all 60 samples. **B**. The distribution of peptides, unique peptides, and identified proteins in this study.

**Supplementary Figure 2 General overview of global proteome profiling**

**A**. Proteomic data filtered at different levels. FDR: false discovery rate, CV: coefficients of variation, IQR: interquartile range, FOT: fraction of total. **B**. Distribution of the protein sequence coverage across all 60 samples from 60 gastric cancer patients. **C**. Distribution of quantified proteins across all 60 samples. **D**. Spearman’s correlation coefficient of the protein ratios in 30 GC tissues and 30 matched healthy tissues. **E**. Distribution of log10(FOT)+7 of the identified proteins in 30 GC tissues and 30 matched healthy tissues. Boxes range from the lower quartile to the upper quartile.

**Supplementary Figure 3 General overview of proteome quantification in AGC**

**A**. Relationship between coefficient of variance (CV) and log10(FOT)+7. When FOT reached more than 10^−5^, overall CV significantly decreased. **B**. Relationship between interquartile range (IQR) and log10(FOT)+7. The increased performance of IQR was discontinued after the FOT reached > 10^−5^. **C**. Relationship between quantitative samples and median CV. When quantitative samples reached > 20, median CV significantly decreased. **D**. PCA analysis showing tumor and matched healthy tissues. PC: principal component.

**Supplementary Figure 4 The AUC values of each protein from the four-protein combination in these datasets**

**A**. The AUC of each protein in Qin’s study (LC-MS/MS). **B**. The AUC of each protein in plasma (ELISA). **C**. The AUC of each protein in tissue microarray.

